# Genome-wide association and Mendelian randomization analysis prioritizes bioactive metabolites with putative causal effects on common diseases

**DOI:** 10.1101/2020.08.01.20166413

**Authors:** Youwen Qin, Guillaume Méric, Tao Long, Jeramie D. Watrous, Stephen Burgess, Aki S. Havulinna, Scott C. Ritchie, Marta Brożyńska, Pekka Jousilahti, Markus Perola, Leo Lahti, Teemu Niiranen, Susan Cheng, Veikko Salomaa, Mohit Jain, Michael Inouye

## Abstract

Bioactive metabolites are central to numerous pathways and disease pathophysiology, yet many bioactive metabolites are still uncharacterized. Here, we quantified bioactive metabolites using untargeted LC-MS plasma metabolomics in two large cohorts (combined N≈9,300) and utilized genome-wide association analysis and Mendelian randomization to uncover genetic loci with roles in bioactive metabolism and prioritize metabolite features for more in-depth characterization. We identified 118 loci associated with levels of 2,319 distinct metabolite features which replicated across cohorts and reached study-wide significance in meta-analysis. Of these loci, 39 were previously not known to be associated with blood metabolites. Loci harboring *SLCO1B1* and *UGT1A* were highly pleiotropic, accounting for >40% of all associations. Two-sample Mendelian randomization found 46 causal effects of 31 metabolite features on at least one of five common diseases. Of these, 15, including leukotriene D4, had protective effects on both coronary heart disease and primary sclerosing cholangitis. We further assessed the association between baseline metabolite features and incident coronary heart disease using 16 years of follow-up health records. This study characterizes the genetic landscape of bioactive metabolite features and their putative causal effects on disease.

## Introduction

The circulating metabolome reflects the compendium of metabolic compounds which operate as inputs, intermediaries and/or outputs for the molecular pathways in blood which sustain or detract from an individual’s health^1^. While many important metabolites have been identified and their role(s) in human diseases characterized, there are thousands, potentially millions, of as yet unidentified metabolites which may be causal or predictive of future health status. Since metabolite identification, the defining of molecular composition and structure, typically requires multiple costly technologies run on valuable biospecimens, there is a need for principled and efficient prioritization of metabolites for targeted, resource-intensive follow-up.

Genome-wide association studies (GWAS) have uncovered hundreds of genetic variants associated with levels of circulating metabolites^2-10^, the vast majority of which are chemically identified. The genes implicated by metabolite-associated genetic variants also show evidence of a functional link between metabolite levels and complex diseases^4^. For instance, the associations between fibrinogen A-α phosphorylation (FAαP) and three genetic loci (*ABO, ALPL* and *FUT2*) support the role of FAαP as a biomarker for acute myocardial infarction, as *ABO* and *ALPL* are also associated with coronary artery disease^4^. Furthermore, Mendelian randomization (MR)^11^ techniques have identified metabolites with evidence of causal effects on disease risk^7^.

Recent advances in untargeted metabolomic profiling, especially liquid chromatography mass-spectrometry (LC-MS), have opened the possibility of detecting and quantifying tens of thousands of discrete chemical features from human blood samples^12^. Our group has pioneered chemical profiling of human plasma using an LC-MS/MS approach specifically developed for profiling of small, polar lipophilic bioactive metabolites in human plasma^13^. These approaches enable detection and relative quantitation of bile acids, sterols, free fatty acids, polyunsaturated fatty acids and oxylipins (e.g. eicosanoids, docosanoids, resolvins, etc), as well as thousands of related metabolites^13,14^. These metabolites are sensed through cell surface and nuclear hormone receptors and serve as critical signaling agents among cellular pathways and systems, mediating a wide variety of processes including host inflammatory response^15-17^, cellular development^18,19^, and nutrient absorption^20,21^. A number of these metabolites have been implicated in the pathobiology of human diseases, including autoimmune disease, cancer, metabolic disease / diabetes, and cardiovascular disease^22-24^. Given their placement in key biological pathways, monitoring of bioactive lipids yields unique insight into the current and future health of monitored individuals and, with likely many yet undiscovered functions of these compounds, will allow for expansion of known biology and discovery of new therapeutic strategies.

While only a minority of bioactive metabolites are chemically identified, their prioritization via biological and clinical utility is under intense investigation. The integration of GWAS, MR and untargeted metabolomics allows for an unbiased, wide-angle approach to prioritize identified and unidentified metabolites, metabolite fragments and metabolic compounds (here termed ‘metabolite features’) for chemical characterization. In the context of common diseases, such an approach may highlight predictive biomarkers and causal metabolic factors which have therapeutic potential.

In this study, we used untargeted high-throughput LC-MS metabolomics to quantify ∼11,000 metabolite features in blood plasma samples of nearly 10,000 individuals with matched genome-wide genotype data. We conducted a discovery GWAS, replication and meta-analysis, then evaluated an extensive set of genetic loci with known and previously unknown roles in metabolism. An ensemble two-sample MR approach was used to assess the predicted causal effects of both identified and unidentified bioactive metabolites on 45 common diseases. We then evaluated the effect of lifetime exposure of bioactive metabolite levels to disease-free survival from baseline using ∼16 years of electronic health record follow-up. Taken together, this study presents a series of findings, including genome-wide maps of bioactive metabolite associations and pleiotropy as well as an unbiased MR-based prioritization of identified and unidentified metabolites with putative causal effects on disease.

## Results

### Genetic variants in 118 loci associate with levels of 2,319 circulating metabolite features

We first performed a discovery genome-wide association scan on the abundances of 11,067 circulating bioactive metabolite features and 7,979,834 genotyped or imputed genetic variants from 7,013 individuals in the population-based FINRISK02 cohort (**Figure 1A; Methods**). Of these metabolite features, 818 were chemically identified metabolites (414 confirmed or putative eicosanoids^11^, 100 free fatty acids (FFA), 66 polar compounds, 47 very-long-chain dicarboxylic acids (VLCDCA), 47 fatty acids esters of hydroxy fatty acids (FAHFA), 35 bile acids, 14 docosanoids, 14 endocannabinoids and 4 sterols). A further 10,249 metabolite features were, as yet, unidentified (**Table S1**). Stringent quality control and normalization procedures were applied to both LC-MS and genetic data (**Methods**). After the univariate genome-wide scan for each bioactive metabolite feature, a joint model was fitted to all genome-wide significant genetic variants to identify a final set of conditionally independent variants reaching significance together with their conditional effect size estimates (**Methods**).

**Figure 1:**
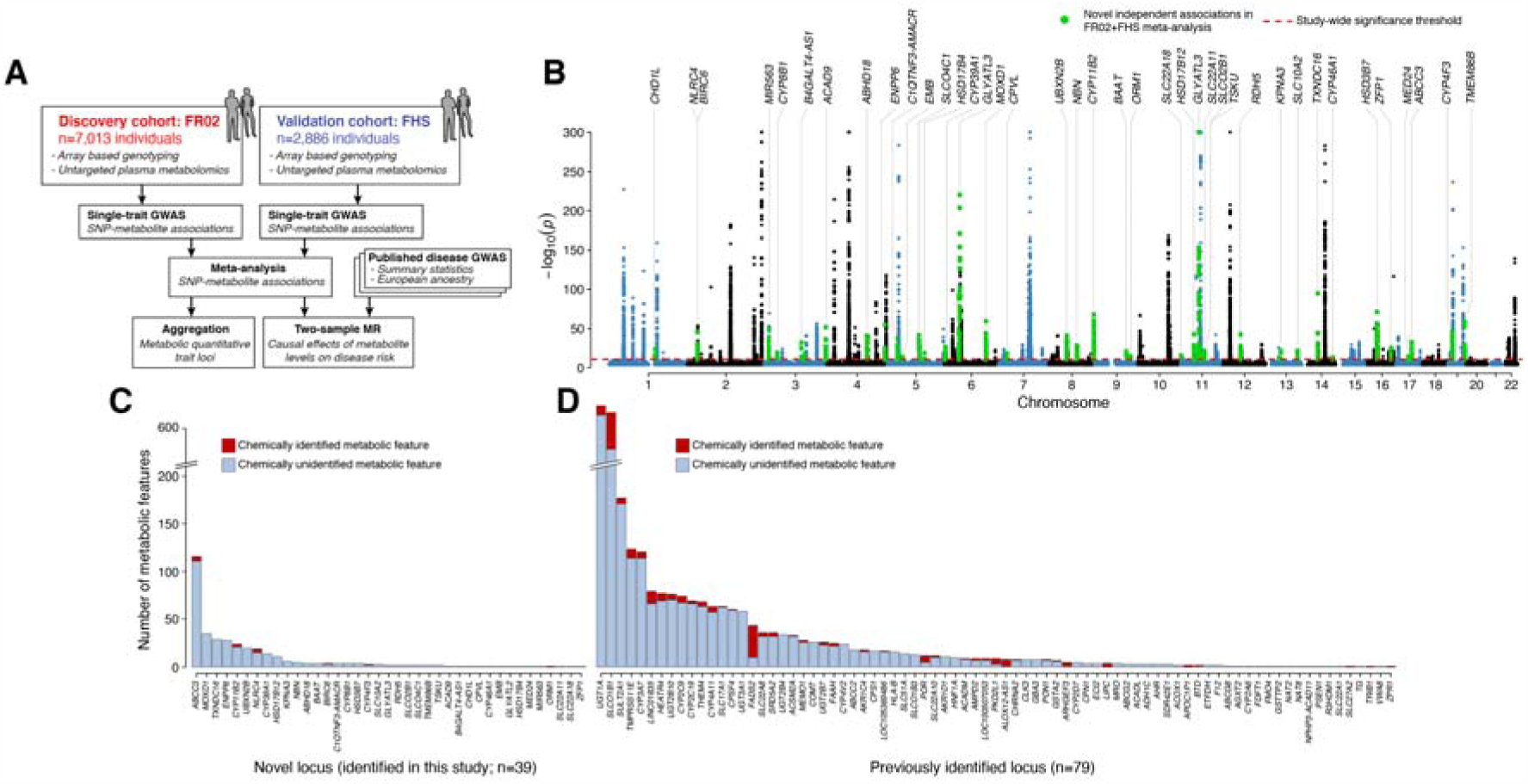
Genome-wide association analysis of circulating metabolite features. (**A**) Summary of the workflow for this study, (**B**) Manhattan plot of associations between genetic variants and circulating metabolite features. Novel loci are highlighted in green and annotated with locus name. The red dashed line indicates study-wide significance (*p*=1.45×10^−11^). For SNPs with multiple associations, only the lowest *p* value is shown. The y-axis is truncated at −log10(*p*)=300 for improved visualization, and SNPs with *p*>10^−4^ are omitted. (**C**) Novel and (**D**) previously reported loci associated with chemically identified (red bars) and chemically unidentified (blue bars) metabolite features. (FHS: Framingham Heart study; FR02: FINRISK02; GWAS: genome-wide association studies; MR: Mendelian randomization.)

A majority of metabolite features (5,874, 57%) had at least one genetic variant reaching genome-wide significance (*p*<5×10^−8^) and were taken forward for replication in the Framingham Heart Study (FHS, N=2,886). Of these associations, the vast majority (4,799) were replicated in FHS at statistical significance (*p*<0.01; **Table S2 & S3**). At a stringent study-wide significance threshold accounting for the number of independent metabolite features (*p*<1.45×10^−11^; **Methods**), 3,602 SNP-metabolite associations were detected in FINRISK02, replicated in FHS at *p*<0.01, and maintained study-wide significance in meta-analysis (**Table S2**). A set of suggestive associations reaching genome-wide but not study-wide significance is given in **Table S3**.

This final set of 3,602 SNP-metabolite associations passing study-wide significance (*p*<1.45×10^−11^) comprised 2,319 metabolite features (161 chemically identified) and 118 genetic loci, of which 39 loci had no previously reported association with blood metabolites (**Figure 1B, Table S4**). Of the 118 genetic loci, 75% were associated with multiple metabolite features, and 31% of metabolite features were associated with multiple genetic loci (**Table S5 & S6**).

Of the 265 associations with chemically identified metabolites, 152 involved eicosanoids (**Table S2**). The strongest associations were between levels of dihydrotestosterone (putative 5α-androstan-17β-ol-3-one) and a variant proximal to *SLC22A8* and *SLC22A24* (rs17713514; meta-analysis *p*<1×10^−300^). *SLC22A24* encodes a solute carrier that has recently been linked to the transport of steroid conjugates^25^, and *SLC22A8* (encoding OAT3) is a recently identified metabolite QTL^9^ that has also been reported in functional experiments to be associated with a wide range of metabolites, including bile acids, flavonoids, nutrients, amino acids and lipids^26^. In addition, levels of dihydrotestosterone were independently associated with variants mapping to *SLC22A9* (rs147394024, meta-analysis *p*=1.2 ×10^−48^) and *SRD5A2* (rs2208532, meta-analysis *p*=7.2 ×10^−26^), which itself encodes 3-oxo-5α-steroid 4-dehydrogenase-2. Furthermore, tetrahydrocortisol levels were associated with variant (rs9994887, meta-analysis *p*=1.2×10^−28^) proximal to *UGT2B15*, encoding UDP-glucuronosyltransferase 2B15, an enzyme involved in the catabolism of xenobiotic compounds and the metabolism of androgens^27^.

Loci previously reported to be associated with blood metabolites showed diverse associations with chemically identified and unidentified metabolite features, and were enriched for pleiotropy as compared to loci without a previous reported metabolite association (*p*<0.0001, Wilcoxon rank-sum test) (**Figure 1C & 1D, Table S5**). Novel loci were associated with 13 identified (including 5 eicosanoids) and 305 unidentified metabolite features (**Table S6**), indicating a sizable component of uncharacterized but genetically controlled metabolism. These findings raised a series of hypotheses.

### Previously unreported loci associated with bioactive metabolites

Two loci *GLYATL2* (11q12.1) and *GLYATL3* (6p12.3) encode glycine N-acyltransferases which showed the strongest associations with 3 chemically unidentified metabolite features (**Figure 1B, Table S6**). Importantly, the *ABCC3* (17q21.33) locus harbored the greatest number of associations with 111 chemically unidentified metabolite features, as well as four identified metabolites (leukotriene D4, putative tauroursodeoxycholic acid, putative 11a-Hydroxyprogesterone b-D-glucuronide, and putative 1,3,5(10)-Estratrien-3,17b-diol diglucosiduronate). *ABCC3* encodes an ATP-binding cassette transporter, multidrug resistance-associated protein 3 (MRP3), known to interact with metabolites through its role as a multidrug exporter, in particular via efflux of potentially toxic endogenous and exogenous compounds from the cell^28,29^. Leukotriene D4 is an endogenous compound derived extracellularly from leukotriene C4, whose cellular release is mediated by MRP3^30,31^. Tauroursodeoxycholic acid is a taurine conjugated compound converted from ursodeoxycholic acid, a secondary bile acid synthesized in liver. Previous studies have revealed that MRP3 is involved in the liver regeneration by pumping out excessive bile acids^32,33^. In addition, mouse study revealed that MRP3 involved in the transport of glucuronidated compounds^34^. 1,3,5(10)-Estratrien-3,17b-diol is a major form of estrogen in human. It has been reported that 17β-Glucuronosyl estradiol is a substrate of MRP3^27,35^ and ethynylestradiol (a synthetic estrogen) increases expression of MRP3 in a rat model^36^. Taken together, our findings support the diverse roles of MRP3 in influencing the systemic metabolism and human health.

Furthermore, *UGT2B7* (UDP-Glucuronosyltransferase-2B7, 4q13.2) was associated with 26 metabolite features, including three identified metabolites (bilirubin, palmitoleic acid, and a putative eicosanoid). These findings consistent with previous studies showing that UDP-glucuronosyltransferase has a diverse set of unrelated endogenous substrates and regulates metabolism^37,38^. This locus was reported to be suggestively associated with plasma metabolites^39^; however the signal in our study was very strong (meta-analysis *p*=1.43×10^−213^).

### *SLCO1B1* and *UGT1A* loci are highly pleiotropic

*UGT1A* (2q37.1) and *SLCO1B1* (12p12.1) showed extensive pleiotropy, with the two loci accounting for a combined 1,495 of the 3,602 total SNP-metabolite associations, and covering 1,211 of the 2,319 metabolite features (**Figure 1D, Table S5 & S6**). Interestingly, only 30 metabolite features (including one eicosanoid) showed associations with both loci. *SLCO1B1* encodes the solute carrier organic anion transporter family member 1B1 (OATP1B1), which is specific to the liver and involved in the transport of various compounds and drugs, including both statins and antibiotics, from the blood into the liver. *SLCO1B1* has been linked to the circulation of several fatty acids as well as statin-induced myopathy^40^, where *SLCO1B1* variants (such as rs4149056) have been linked to statin response and risk of heart failure^41^. Notably, we found that levels of leukotriene D4, a vasoconstrictor eicosanoid, were strongly independently associated with two SNPs at the *SLCO1B1* locus (rs4149056, meta-analysis *p*=5.4×10^−193^; rs11045856, meta-analysis *p*=7.15×10^−108^; **Table S2**). Rs4149056 showed diverse associations with other eicosanoids, palmitoleic acid, and unidentified metabolite features (**Table S2**). *UGT1A*, encoding UDP glucuronosyltransferase 1 family polypeptide A cluster, is a complex locus of several UDP-glucuronosyltransferases involved in glucuronidation and metabolism. This family of UDP-glucuronosyltransferases are assembled via differential splicing of numerous exons^42,43^. In previous GWASs, variants in *UGT1A* have been shown to be associated with multiple circulating metabolites, including bile pigments^3,4,6,10^. Our findings were consistent with this and indicated a systemic metabolite role for *UGT1A*.

### Cytochrome P450 loci

Cytochrome P450 (CYP) enzymes are known to have diverse roles in the metabolism of both endogenous and exogenous compounds, including drugs, eicosanoids, bile acids etc^44^. In total, 12 loci encoding seven CYP families were associated with 316 metabolite features in meta-analysis, collectively accounting for 12% (433/3602) of total SNP-metabolite associations (**Table S5 & S6**). Notably, the *CYP3A* subfamily locus (7q22.1) was associated with the largest number of metabolite features, including 6 identified and 114 chemically unidentified. The *CYP3A* subfamily is expressed in liver and gut and is known to metabolize >120 commonly prescribed drugs^44^. In addition, two loci *CYP2C19* and *CYP2C9* (10q23.33) encoding the *CYP2C* enzyme family were together associated with over 113 metabolite features (including 5 identified metabolites) (**Table S6**).

### Putative causal effects of metabolite features on disease

To prioritize metabolite features using their evidence for causal effects on disease, we utilized two-sample MR together with GWAS summary statistics for 45 common diseases (**Table S7**). An ensemble of five different MR methods were used, and directionally-consistent statistical significance from at least three methods was necessary to infer causality (**Methods**). Given the frequency of pleiotropic effects, we also used a second step correction using MR-PRESSO^45^ to control for horizontal pleiotropic outliers. At least five genetic instruments were required for a metabolite feature to be considered for causal inference (**Methods, Table S8**). Of the 70 metabolite features meeting these criteria, 31 showed evidence of causal effect on at least one disease (**Figure 2, Table S9**), and 30 of these were robust after further correction with MR-PRESSO. The majority of metabolite features with causal effects were for coronary heart disease (CHD) and primary sclerosing cholangitis (PSC), with one metabolite feature each affecting risk of schizophrenia, bipolar disorder or rheumatoid arthritis. Amongst the 31 metabolite feature levels, there were a wide range of correlations with most exhibiting weak positive correlations as well as ∼5 clusters of features whose levels were in moderate-high correlation.

**Figure 2.**
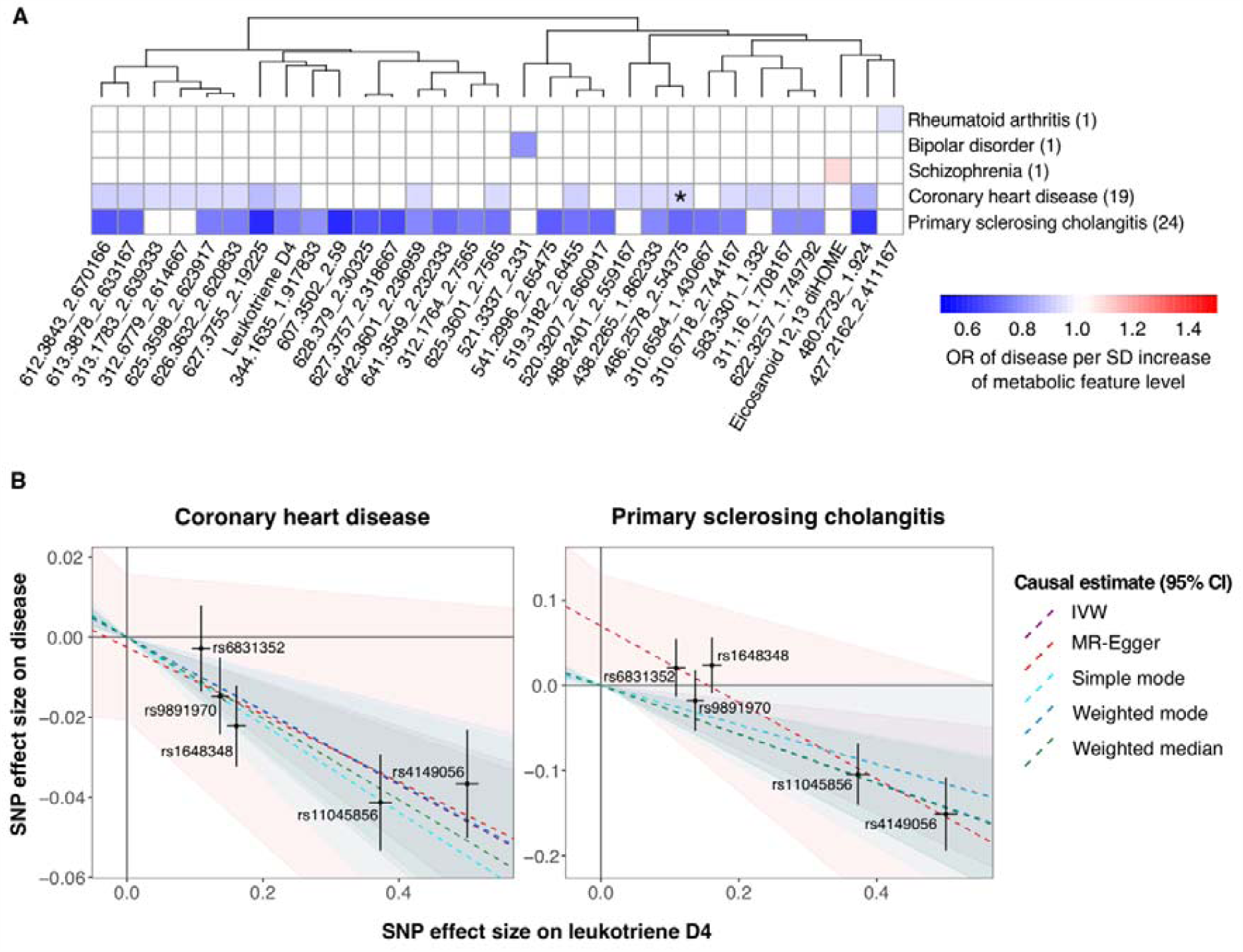
Causal effects of circulating metabolite features on common diseases. (**A**) Predicted causal (red) and protective (blue) effects reaching statistical significance in three of the five MR methods tested. Causal effect estimates are from the weighted median method. Asterisk (*) indicates causal effect which was not significant in MR-PRESSO outlier-corrected test. (**B**) Dose-response plots of leukotriene D4 (LTD4) levels on coronary heart disease and primary sclerosing cholangitis. Causal estimates of the five MR methods are shown with confidence intervals indicated by shaded area in corresponding color.

Of the 15 metabolite features with putative causal effects on both PSC and CHD, all were inverse associations: lifetime exposure to low metabolite feature levels increased risk of CHD and PSC, with the latter exhibiting a somewhat greater effect size (**Figure 2A**). Lifetime exposure to elevated leukotriene D4, a basophil-secreted metabolite known to be involved in the induction of smooth muscle contraction, vasoconstriction and vascular permeability^46^, was predicted to reduce risk of both PSC and CHD (**Figure 2B**). While evidence of a pleiotropic effect (MR-Egger intercept=0.07, *p*=0.02) for PSC was observed, the causal estimates were consistent across all testing methods. Leukotriene D4 was only weakly correlated with other unidentified metabolite features (**Figure S2**), with metabolite IDs 627.3755_2.19225 (*m/z* ratio 627.3755), 480.2732_1.924, and 612.3843_2.670166 showing similar or stronger causal effect sizes on risks of CHD and PSC (**Figure 2A**).

### Association of metabolite features with incident coronary heart disease

Using the baseline bioactive metabolite features, we next assessed CHD-free survival for incident CHD via the linked EHR data available in both cohorts (**Methods**). While PSC is relatively rare in the population (with only 9 incident cases in FINRISK02), we were powered to detect baseline metabolite feature associations with CHD (541 and 97 incident CHD cases in FINRISK02 and FHS, respectively). Time-to-event Cox proportional hazards models were used in both cohorts and then meta-analyzed (**Methods**).

Of the 19 metabolite features with putative causal effects on CHD, six were associated with incident CHD at FDR-adjusted significance (**Table S10, Figure 3**), with an additional metabolite feature associated at nominal significance (*p*<0.05). For these seven metabolite features, there was opposing direction of effect for the MR-based lifetime exposure estimate and time-to-event Cox model estimate, with the former being negative and the latter being positive (**Figure 3**). The corresponding hazards ratios from the Cox models of each metabolite feature in FINRISK02 and FHS were consistently positive in both cohorts (**Table S10**).

**Figure 3.**
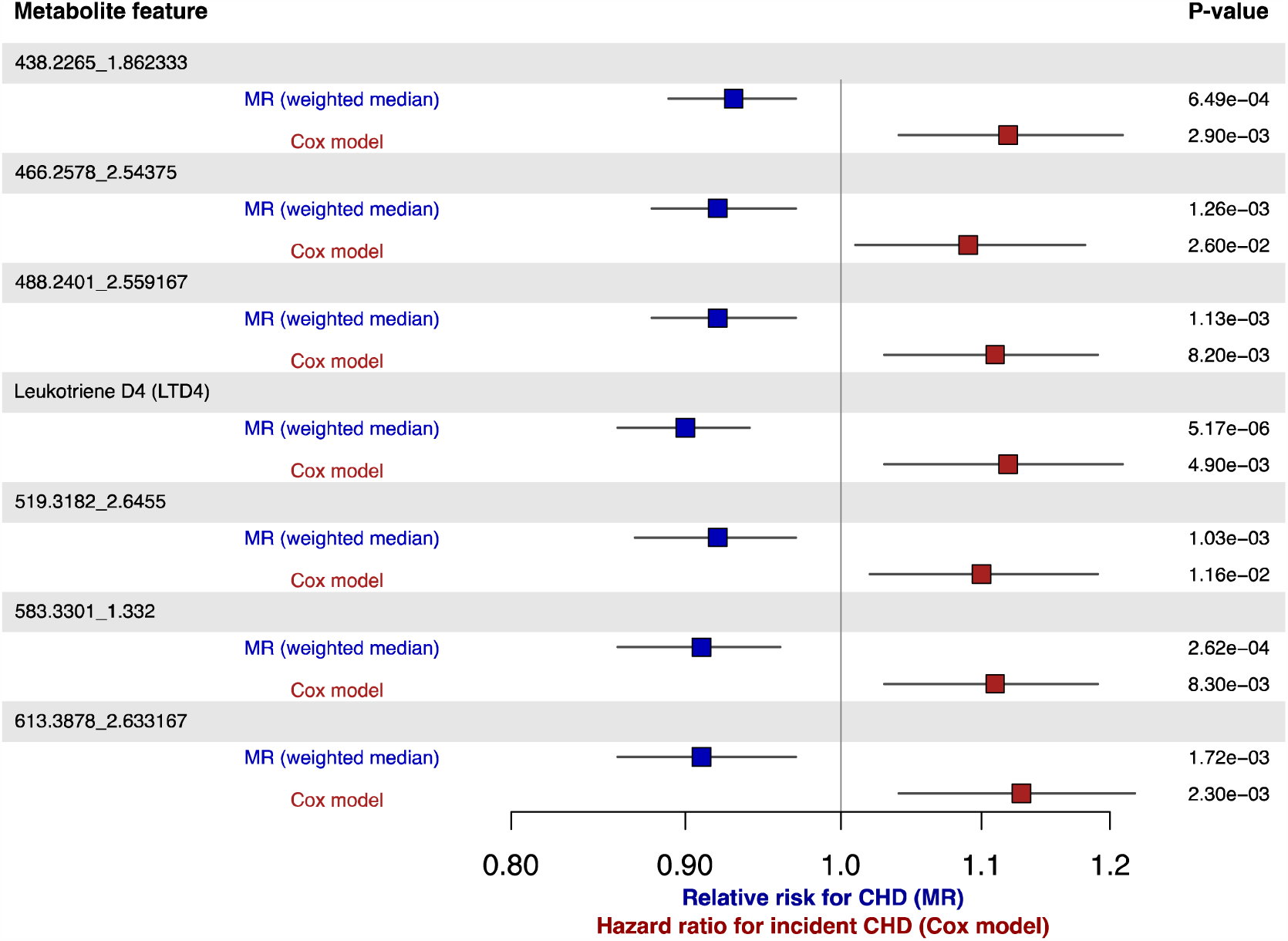
Comparison of MR-based causal effects and association of baseline metabolite feature levels with incident coronary heart disease risk. A forest plot of MR-based effect sizes (weighted median method) and hazard ratios were meta-analysis of Cox regression in FINRISK02 and FHS. Effect sizes are per SD of metabolite feature.

## Discussion

In this study, we investigated the genetic associations of over 10,000 bioactive metabolite features and their relationships with common diseases. We identified 118 genetic loci harboring variants robustly associated with the levels of 2,319 metabolite features, 91% of which were chemically unidentified compounds, suggesting a largely unexplored reservoir of genetic control of the circulating metabolome. We identified 39 genetic loci previously unlinked to blood metabolites and highlighted loci with extensive pleiotropy for bioactive metabolites. We found causal effects for multiple identified and unidentified bioactive metabolites on diverse common diseases, and investigated the baseline relationship of putatively causal metabolite features with incident coronary heart disease.

Our findings were consistent with known metabolic pathways and indicate potentially new gene functions. First, membrane transporters play important role in homeostasis by regulating the transcellular movement of solutes between body fluid compartments. The communication of small-molecule substrates between cells requires the activity of both SLC (generally influx) and ABC (efflux) transporters^47^, of which SLC22A8 and SLCO1B1 are two major transporters with particular relevance to drug compounds^47^. Loci encoding *SLC22A8* and *SLCO1B1* were associated with diverse metabolite features, indicating broad substrate specificity. Second, although the role of ABCC3 (MRP3) in metabolism has been established^28,47^, to our knowledge genetic associations have not been previously reported. *ABCC3*, a hepatocyte efflux pump for bilirubin, was associated with four identified bioactive metabolites with previously known relationships to MRP3 as well as over 100 unidentified metabolite features which may be part of these pathways or lead to novel metabolic roles for MRP3. Third, our findings suggest the *UGT1A* and *SLCO1B1* loci have central roles in bioactive metabolic pathways, accounting for ∼40% of our study-wide significant associations. Given the splicing complexity of the *UGT1A* locus and its propensity to encode a diversity of UDP-glucuronosyltransferases, this may indicate genetic control of exon usage and downstream enzymatic functions.

The combination of untargeted metabolomics, GWAS and MR detected a set of 31 metabolite features, most of which as yet are unidentified, which showed causal effects on disease risk and which now may be prioritized for further investigation. These included 15 bioactive metabolite features with shared causal effects on both CHD and PSC. While there has been little to link between CHD and PSC in the existing literature. Our findings indicate that bioactive metabolites, including leukotriene D4, may comprise previously unknown causal metabolic pathways modulating risk of both diseases. As a chronic autoimmune disease, PSC is a progressive disease mainly associated with the hepatic system and inflammation of the bile ducts, consistent with putative causal effects of leukotriene D4, an inflammatory mediator known to be released by basophils^48^. We would speculate that other bioactive metabolites amongst the 15 affecting CHD and PSC risk are also mediators of inflammation, a broad but recognized causal process underlying both diseases.

Notably, for bioactive metabolites associated with CHD, we found directional inconsistency between baseline risk prediction models and risk conferred by lifetime exposure (derived from MR). This adds to previous evidence for significant but directionally opposed biomolecular associations with cardiovascular diseases in MR and observational analyses. A recent proteome analysis found that genetic predisposition to higher plasma MMP-12 levels was predicted to reduce risk of coronary disease and atherosclerotic stroke, despite observational studies finding a positive association with cardiovascular disease risk^49^. A separate study found levels of PON1, a major anti-atherosclerotic component of high-density lipoprotein (HDL), to be negatively associated with CHD in observational analyses but with MR results predicting higher PON1 to increase risk of CHD^50^. Authors hypothesized that the CHD condition may be linked to a downregulation of PON1 resulting in lower plasma proteins.

These findings appear to be robust yet puzzling. We hypothesize several scenarios which may explain these data. First, the causal effect estimated using MR is thought to indicate a lifetime average effect^51,52^; however, biological pathways are highly dynamic and biomolecules may have age or time-varying effects. Second, sub-clinical atherosclerosis or other relevant disease states may rewire biochemical and metabolic pathways such that disease risk is increased but normal biological functions (as estimated by MR) are compromised. Third, MR approaches assume linearity for the examined causal effect and for the genetic effects on the exposure and the outcome^53^. However, biological risk factors may have non-linear effects on the outcome, For example, high body mass index (BMI) is a risk factor for type 2 diabetes (T2D) in adults; however, high BMI can be protective in infants^54^ and early childhood famine is associated with higher T2D risk in later life^55^.

In conclusion, this study shows the efficiency of coupling untargeted metabolomics, GWAS and MR to prioritize the bioactive molecular features with causal effects on disease. In doing so, it highlights loci harboring potentially key enzymes, uncovers new insights into bioactive molecular pathways, and raises salient questions of observational effects and MR-based lifetime effects of molecular exposures which may have important therapeutic implications.

## Materials and Methods

### Study cohorts

FINRISK is a nation-wide population-based study of Finland periodically recruiting participants every 5 years since 1972^56^. Participants included in this study were randomly recruited from six defined geographical areas across Finland in 2002^57^. Blood samples were taken during their visits. The follow-up data until 2018 were extracted from Finnish national hospital discharge registries, drug reimbursement registries and causes-of-death registries. In summary, there were 8,738 individuals, 4,688 (54%) females and 4,050 (46%) males, with baseline age between 24 to 75 years.

The Framingham Heart Study (FHS) is a multi-generational population-based study. The data used in this study was from the FHS offspring exam 8 participants^58^. This subset of the FHS cohort included 2,886 individuals, with average age of 66.4 (SD=9.0) years old, average BMI of 28.3 (SD=5.4) kg/m^2^, and 54% were female.

The FINRISK 2002 survey has been approved by the Ethical Committee on Epidemiology and Public Health of the Helsinki and Uusimaa Hospital District (decision number 87/2001) and the participants have provided an informed consent. The study is conducted according to the World Medical Association’s Declaration of Helsinki on ethical principles. The FHS study data was accessed via dbGaP (approved study 2014.2023).

### Metabolomic profiling with liquid chromatography mass spectrometry

Plasma samples were randomly assigned to 90 plates and measured by LC-MS (Thermo Vanquish UPLC coupled to a Thermo Q Exactive Orbitrap mass spectrometer) in a randomized order. Batch effects and machinery/chemical variance were assessed by 19 spike-in internal standards in each measured sample and three pooled plasma control samples per 96-well plate. In-house R scripts were used to process the data including initial bulk feature alignment, MS1-MS3 data parsing, pseudo DIA-to-DDA MS2 deconvolution, and CSV-to-MGF file generation. Subsequently, mzMine 2.21^59^ was employed for feature extraction, secondary alignment and compound identification.

Metabolite features were filtered if they met any of the conditions: (1) less than 5 observations per plate, (2) a difference in observation missingness of 50% or greater from the median plate missingness, (3) a plate standard deviation that differed from the median plate standard deviation by a factor of 2.5 or greater, (4) a relative median offset of 4 or greater, and (5) overall missingness >50%. This filtering resulted in an intensity profile of 11,067 measured metabolite features across 8,291 samples.

For FINRISK02, a two-step normalization was applied to the intensity data to control for technical variation. The first step was to remove variation observed in external bracket pooled plasma with RUV^60^, a tool implemented in R package *MetNorm* to remove unwanted variation of metabolomics data. The second step was to remove variation observed in internal standards with RUV. There were two components for each RUV process, RUV-random (function *NormalizeRUVRand*) and RUV-Kmeans (function *NormalizeRUVRandClust*). RUV-random was to remove *k* technical factors estimated from external pooled plasma or internal standards, where *k* was set to the number of principal components explaining >97.5% of variation. RUV-Kmeans was used to refine the *k* technical factors to better represent the biological variation. Following RUV, the plate median was subtracted from the resulted matrix to get normalized matrix. Samples which exhibited excessive missing data of >10% were removed (N=449), and K-nearest neighbors (R package *impute*)^61^ was used to impute missing data into the remaining samples. Post-QC normalized metabolite feature data was standardized to mean 0 and standard deviation of 1. The final intensity matrix consisted of 11,067 metabolite features and 7,842 samples.

For FHS, normalization of the metabolite intensity values included a four-step process: (1) shift to positive values, (2) cap outliers, (3) log transformation, and (4) scale to a normal distribution. Metabolite features presented in >20% individuals were kept for the analysis.

### Genotyping, imputation and quality control

For FINRISK02, genotyping was performed on Illumina genome-wide SNP arrays (the HumanCoreExome BeadChip, the Human610-Quad BeadChip and the HumanOmniExpress) and has been described previously^62,63^. Stringent criteria were applied to remove samples and variants of low quality. Samples with call rate <95%, sex discrepancies, excess heterozygosity and non-European ancestry were excluded. Variants with call rate <98%, deviation from Hardy-Weinberg Equilibrium (*p*<1×10^−6^), and minor allele count < 3 were filtered. Data was pre-phased by using Eagle2 v2.3^64^. Imputation was performed using IMPUTE2 v2.3.0^65^ with two Finnish-population-specific reference panels: 2,690 high-coverage whole-genome sequencing and 5,092 whole-exome sequencing samples. To evaluate the imputation quality, we compared the sample allele frequencies with reference populations and examined imputation quality (INFO scores) distributions. Imputed SNPs with INFO >0.7 were kept for analysis.

Post imputation quality control was carried out by using plink v2.0^66^. Samples with >10% missing rate were removed. Individuals with extreme height or BMI values were further excluded (31 individuals with height<1.47m; 5 with BMI >50 were removed). We also removed 42 pregnant women since pregnancy is known to have dramatic changes to body metabolism. Both genotyped and imputed SNPs were kept for analysis if they met the following criteria: call rate >90%, no significant deviation from Hardy-Weinberg Equilibrium (*p*>1.0×10^−6^), and minor allele frequency >1%. The post-QC dataset comprised 7,013 individuals and 7,980,477 SNPs. SNPs with ambiguous A/T or C/G alleles were removed in meta-analysis.

FHS genotyping was performed using “Affymetrix Nsp, Sty and 50K gene centric” arrays. To impute genotypes for SNPs, short insertions and deletions (indels), and larger deletions that were not genotyped directly but are available from the 1000 Genomes Project, imputation of 8,493,311 genetic variants was performed with Minimac3 using the 1000 Genomes Project Phase I Integrated Release Version 3 Haplotypes (2010-11 data freeze, 2012-03-14 haplotypes).

### Genome-wide association analysis

Univariate testing was performed using BOLT-LMM (v2.3.2)^67^, a Bayesian mixed model. The kinship matrix was constructed using 106,201 SNPs selected by pruning the hard-called SNPs with r^2^ < 0.1 (plink2 command --*indep-pairwise 1000 80 0*.*1*). Genetic principal components were calculated using FlashPCA2^68^ on the pruned SNPs. Leave-one-chromosome-out (LOCO) analysis within BOLT-LMM was used to avoid proximal contamination. The linear mixed model included the following covariates for FINRISK02: genotyping batch, age, gender and top 10 genetic principal components; for FHS, the covariates were age and sex. The average genomic inflation factor was 1.0067 (range 0.9776 to 1.0389) and a positive correlation (Pearson correlation coefficient was 0.57) was observed between the genomic inflation factor and SNP-heritability (**Figure S1**). For those SNPs reaching genome-wide significance (*p*<5×10^−8^), to obtain conditionally independent SNP-metabolite associations GCTA-COJO^69^ was used to conduct step-wise conditional and joint analysis on individual genotype data. Given the large number of metabolite features, the number of effective tests was based on eigenvalue variance and estimated using matSpDlite^70,71^. For all 11,067 metabolite features, the number of effective tests was 3,450 thus the study-wide significant threshold for SNP-metabolite associations was *p*<1.45×10^−11^.

Associations passing genome-wide significance in FINRISK02 were taken forward for validation and meta-analysis in FHS, with the mapping of metabolite features between the two cohorts being based on MS1 and MS2 spectra alignment of metabolites with similar mass charge ratio and retention time in principle^72^. Of the metabolite features, 76% (4474/5874) were matched to at least one feature present in >20% people in the FHS cohort. For metabolites with multiple matches, the strongest associations (lowest *p* values) were kept. Meta-analysis was performed using the inverse variance weighted method for fixed effects (R package *meta*). Only SNP-metabolite associations reached *p*<5×10^−8^ in discovery cohort and *p*<0.01 in replication cohort were meta-analyzed.

ANNOVAR^73^ was used to annotate significant SNPs. SNPs were assigned to genetic loci using the 200kb region flanking the top SNPs (i.e. lowest *p* value)^74^. This aggregation process started from the overall top SNP, followed by the second top SNP of the remaining SNPs and so on, until there was no SNP left. Loci names were determined from the nearest genes to the associated top SNPs. No merging was performed for neighboring loci, a particular SNP could be assigned to two different loci if those were both located within 200kb of the SNP position. As such, 41 SNPs in total were assigned to more than one locus; in these cases, we reported both loci in **Table S4** but do not count them twice when summarizing results. A locus was defined as novel if it was not located within 200kb of any previously reported variants associated with blood metabolites in the GWAS Catalog repository (release 2020-01-27), or in the latest table from Kastenmüller et al.^75^ (http://www.metabolomix.com/list-of-all-published-gwas-with-metabolomics; last accessed 02/2020).

### Mendelian randomization

For genetic instruments, we utilized SNPs reaching genome-wide significance in meta-analysis. Summary statistics of SNP-disease associations were extracted from MR-base database using R package *TwoSampleMR*^76^. If an index SNP was not present, the strongest proxy SNP was used or set to missing if r^2^<0.8). GWAS summary statistics were required to include European ancestries and be based on at least 1,000 individuals (**Table S7**). For each metabolite-disease analysis, SNPs were clumped using a linkage-disequilibrium threshold of r^2^<0.05 in a 500kb window to minimize the impact of correlated SNPs on causal estimates. As MR analysis with multiple instruments is more reliable, five or more genetic instruments were required for a metabolite to be taken forward for MR analysis. The effect allele was taken to be the effect-increasing allele of metabolite in FINRISK02. We estimated causal effects using an ensemble of five widely utilized methods: inverse variance weighted (IVW)^77^, simple mode^78^, weighted mode^78^, weighted median^79^, and MR-Egger^80^. As these methods have different assumptions, agreement among multiple methods would indicate a robust estimate of causal effects^81^. We defined a significant causal effect as *p*<0.05 in three of the five selected methods. For significant causal estimates, details of genetic instruments are provided in **Table S8**. As a second step, for putative causal associations passing three of the five methods in the ensemble, we then used MR-PRESSO^45^ to detect and correct for horizontal pleiotropic outliers.

### Cox proportional hazards models

To test the association between metabolite feature and incident CHD in both FINRISK02 (16 years follow-up; 541 incident events) and FHS (6 years follow-up; 97 incident events), Cox proportional hazards regression (*coxph* function in the *survival* R package) was utilized to predict the CHD event for metabolite feature. Metabolite levels were at log10 scale. Covariates included age, sex, and log-transformed BMI. Participants with prevalent CHD at baseline were excluded. Cox models were sex-stratified with time-on-study as the time scale. Fixed effect meta-analysis was conducted to combine the summary statistics from both cohorts. Sensitivity analysis was conducted to ensure associations were robust to LDL-cholesterol, smoking status, hypertension, diabetes as well as medications for lipid-lowering, anti-hypertension and diabetes.

## Data Availability

The data used in this study is available with an approved study proposal via the THL Biobank (FINRISK) and NIH dbGaP (Framingham Heart Study).

## Acknowledgements

The FINRISK02 cohort was mainly funded by budgetary funds of the Finnish Institute for Health and Welfare. Important additional funding has been obtained from the Finnish Academy and domestic non-profit foundations. SCR was funded by the UK National Institute for Health Research (NIHR) (Cambridge Biomedical Research Centre at the Cambridge University Hospitals NHS Foundation Trust). LL is supported by the Academy of Finland (grant n:o 295741). TN is supported by the Academy of Finland (grant n:o 321351), the Finnish Medical Foundation, the Paavo Nurmi Foundation, and the Emil Aaltonen Foundation. VS has been supported by the Finnish Foundation for Cardiovascular Research. MI was supported by the Munz Chair of Cardiovascular Prediction and Prevention. This study was supported by the Victorian Government’s Operational Infrastructure Support (OIS) program.

This work was supported by Health Data Research UK, which is funded by the UK Medical Research Council, Engineering and Physical Sciences Research Council, Economic and Social Research Council, Department of Health and Social Care (England), Chief Scientist Office of the Scottish Government Health and Social Care Directorates, Health and Social Care Research and Development Division (Welsh Government), Public Health Agency (Northern Ireland), British Heart Foundation and Wellcome.

The funders had no role in study design, data collection and analysis, decision to publish, or preparation of the manuscript. The views expressed in this manuscript are those of the authors and not necessarily those of the NHS, the NIHR or the Department of Health and Social Care.

